# COVIDCare@Home: Lessons from a Family Medicine Led Remote Monitoring Program

**DOI:** 10.1101/2020.07.20.20158386

**Authors:** Payal Agarwal, Geetha Mukerji, Celia Laur, Shivani Chandra, Nick Pimlott, Ruth Heisey, Rebecca Stovel, Elaine Goulbourne, R. Sacha Bhatia, Onil Bhattacharyya, Danielle Martin

## Abstract

**Background:** Virtual care for patients with COVID-19 allows providers to monitor COVID-19 positive patients with variable trajectories while reducing the risk of transmission to others and managing healthcare capacity in acute care facilities.

**Objective:** To develop and test the feasibility of a family medicine-led remote monitoring model of care (COVIDCare@Home program) to manage patients with COVID-19 in the community.

**Methods:** This multi-faceted, family medicine-led, interprofessional team-based remote monitoring program was developed at Women’s College Hospital in Toronto, Ontario. A cross-sectional chart review of the first cohort of patients was conducted and learnings from the implementation of CovidCare@Home are described.

**Results:** During the study period, April 8 to May 11, 2020, there were 97 patients (average age 48.6, 62% female) with 424 recorded virtual visits with a median virtual length of stay of 8 days (IQR 5). 5.2% required escalation to an in-person visit with no patients requiring hospitalization. 16% of patients required support with mental and social health needs.

**Interpretations:** A family medicine-led, team-based remote monitoring program can safely be used to manage outpatients diagnosed with COVID-19. Attention to mental and social health needs is critical for this population. Future efforts should consider how to design programs to best support populations disproportionately impacted by COVID-19, something which primary care is well-positioned to do. Further analysis will describe the effectiveness, impact, and satisfaction with the program among patients and providers.

## Background

The COVID-19 pandemic has spread to 213 countries worldwide. Canada has had 107 590 cases and 8783 deaths, as of July 12, 2020 (1). As part of the health system’s response in Ontario, in-person visits were scaled back dramatically; virtual visits became the standard for most non-essential and much essential care. For patients infected with COVID-19, using virtual care to monitor the disease at home allows providers to address patient needs while reducing the risk of transmission to other patients or providers (2–4). The majority of COVID-19 positive patients can safely convalesce at home, but roughly 10% will require hospital admission (5–7). Importantly, some presentations require early identification and acute care treatment to prevent poor outcomes (8–10)], while frequent virtual touch points may reduce unnecessary Emergency Department visits for others (2,6,11)

Ideally, patients with COVID-19 would be supported by their own primary care provider (PCP), with whom they have an established relationship, but many Ontario PCPs reduced services during the first wave of the pandemic or did not have sufficient or reliable infrastructure to support their patients. Lack of comfort managing patients with a novel infectious disease at a distance may also have made primary care management more challenging (2,12). Furthermore, there is no consensus on the optimal model of remote monitoring for COVID-19. Some models are specialty-based (6), while others include both primary care and specialty care physicians (5,13,14). The resource-intensive nature of these models calls into question their sustainability and generalizability. Many models are disease-focused, algorithm-dependent and are not designed to manage patient comorbidities and psychosocial issues that arise during the illness (5,6,13,14).

A family medicine-led interprofessional model of remote home monitoring for COVID positive patients was developed, with a focus on those who did not have a tight connection to primary care. This study describes the model of care and discusses its safety and feasibility in the first five weeks.

## Methods

### Setting

Women’s College Hospital (WCH) is an ambulatory academic hospital located in Toronto, Canada. In late March 2020, the hospital partnered with the Department of Family and Community Medicine at the University of Toronto, and Mount Sinai Hospital (MSH), an acute care academic hospital and part of the Sinai Health System (SHS), to develop a model to care for patients with COVID-19 in the community. The program was operational by April 8, 2020. This study includes a cross sectional chart review of all patients who had their first appointment from the start of the program until May 11, 2020.

### Overall Care Model

COVIDCare@Home was established using the principles and protocol described by Greenhalgh et al. (2) adapted with input from multiple stakeholders based on available evidence (**Supplement 1**). The program offers remote monitoring, using telephone or video visits, 7-days a week by an interprofessional, family medicine led team. Patients also had access to a dedicated on-call service 24-hours a day. Pulse oximeters and thermometers were couriered to patients felt to be at high risk, based on age, comorbid illness and respiratory symptoms. Typically, the program aimed to follow patients from time of referral up to 14 days from symptom onset or, for asymptomatic patients, the date of a positive swab.

### Patient Population

All patients in the Greater Toronto area diagnosed with COVID-19 (swab + or presumed positive) were eligible for the program. Multiple referral pathways were built to support broad access. These included COVID-19 assessment centres at WCH and MSH, the emergency department of MSH, post-discharge from acute care or in-patient rehabilitation services at SHS and directly from primary care providers in the community.

### Care Team

The interprofessional team included a family physician, a family medicine resident, registered nurse (RN), a mental health/social worker, nurse practitioner (NP), and a pharmacist available 7-days a week. Regular video visits were conducted by the family medicine resident or RN depending on patient complexity. Specialists and sub-specialists, including general internal medicine, respirology and psychiatry were available for virtual consults as needed. Given the complexity and uncertainty in treating COVID-19, there were daily huddles at the end of the clinic with team members to review all cases and weekly rounds with the full team and specialists to discuss challenging cases or frequently occurring clinical questions.

### Digital Tools

Care was charted using the EPIC Electronic Medical Record (EMR) at WCH which enables secure, EMR-integrated video visits via Zoom and bi-directional messaging using a patient portal. Patients could participate in video visits using a cell phone, tablet or computer. Care was also provided by telephone when the patient preferred or was unable to connect via video. A website with resources for patients and physicians was developed to facilitate care. A dashboard cataloguing each patient in the program with their risk level for deterioration and active care issues was developed to facilitate daily team huddles. A telephone translation service was used for patients more comfortable in a language other than English.

### Clinical Processes

Initial assessments of all patients were done by the resident supervised by the staff physician. All patients were triaged to low, moderate or high risk using clinical judgement based on: 1) age and comorbidities; 2) trajectory in disease course as patients are more likely to decompensate day 5-12 post symptom onset (15); 3) current symptoms and oxygen saturation and temperature when available; and 4) additional social complexities. Follow-up virtual visits were booked with the resident or RN every 1-3 days based on risk. Risk was reassessed at each visit to inform the care plan and monitoring schedule.

The appropriate team member was notified if patients required additional support and that provider conducted a separate virtual appointment. Nurse Practitioners supported case management of complex patients; Social Workers addressed mental health concerns and provided brief mental health counselling along with supporting access to community resources. Patients’ relationship to a PCP was explored early and efforts made to contact the PCP to facilitate a shared care approach as appropriate with a clear, well communicated discharge plan.

There were several options to escalate care when necessary tailored to the individual patients’ needs and disease trajectory. For medically stable patients who required further workup of symptoms or comorbidities, an in-person visit could be arranged in the Acute Ambulatory Care Unit (AACU), a short stay medical unit at WCH, with a general internist and access to urgent labs and imaging. Acutely ill patients were sent to the emergency department (ED), while home care could be arranged for those who did not want to be transferred to an acute care facility based on their goals of care.

### Data Collection and Analysis

Clinical and contextual information regarding patients and COVID-19 diagnosis was collected during virtual clinical encounters and entered into EPIC using a standardized electronic flowsheet. All data from the flow sheet, basic patient demographics, and program utilization data was electronically extracted. Two research coordinators reviewed patient charts to extract additional information not captured in the flow sheets. Data discrepancies or concerns were reviewed by the study lead (PA) and consensus was reached as a group. Descriptive statistics were used to describe patients and patterns of service utilization.

### Ethics

This study was completed by the investigators without the influence of any commercial sponsor. The study was approved by the local research ethics board at Women’s College Hospital (2020-0058-E).

## Results

### Baseline demographic and clinical characteristics

Baseline demographics and clinical characteristics of patients in COVIDCare@Home are presented in **Table 1**. Ninety-eight patients met the inclusion criteria; one was not included as they did not complete their initial appointment. The mean age was 43.6 years (SD 14.2), with a 1:2 male to female patient ratio. Of 97 patients, 77% had access to a primary care provider. Over half (50.5%) had at least one comorbidity, with 11% identifying three or more comorbidities. Most patients were positive for COVID-19 (88% swab positive and 4.1% presumed), but 5 patients (5.1%) were found unlikely to be infected with COVID-19 over their time in the clinic and their symptoms were attributed to other health conditions.

**Table 1:** Characteristics of patients enrolled in COVIDCare@Home program.

|  | Number (%) of patients | Standard deviation |
| --- | --- | --- |
| **Total patients enrolled** | 97 |  |
| **Age** |  |  |
| *Mean age* | 43.8 years | 14.2 |
| Under age 18 | 1 (1.0%) |  |
| Over age 60 | 17 (17.311%) |  |
| **Sex** |  |  |
| Female | 65 (67.0%) |  |
| Male | 32 (33.0%) |  |
| **Has a Primary Care Provider** | 75 (77.3%) |  |
| **Co-morbidities** |  |  |
| *Number of patients with one or more comorbidities* | 49 (50.5%) |  |
| Asthma | 11 (11.3%) |  |
| Autoimmune/Immunosuppressed | 6 (6.2%) |  |
| CHF | 1 (1.0%) |  |
| CKD | 1 (1.0%) |  |
| Liver disease | 0 (0%) |  |
| COPD | 1 (1.0%) |  |
| Cardiovascular disease | 2 (2.1%) |  |
| Diabetes | 6 (6.2%) |  |
| Hypertension | 11 (11.3%) |  |
| Malignancy | 0 (0%) |  |
| Anxiety | 9 (9.3%) |  |
| Depression | 3 (3.1%) |  |
| Dyslipidemia | 10 (10.36%) |  |
| Other comorbidities | 22 (22.6%) |  |
| **Other Factors** |  |  |
| Alcohol use | 0 (0%) |  |
| Smoking | 5 (5.2%) |  |
| Pregnancy | 4 (4.1%) |  |
| **COVID Status:** |  |  |
| Swabbed positive | 88 (90.7%) |  |
| Presumed positive | 4 (4.1%) |  |
| COVID-19 negative | 5 (5.1%) |  |
| **COVID Transmission Risk Factors:** |  |  |
| Occupation | 55 (56.7%) |  |
| Long-term care home | 18 (18.6%) |  |
| Acute Care | 9 (9.3%) |  |
| Shelter | 9 (9.3%) |  |
| Complex Continuing Care | 12 (12.4%) |  |
| Grocery Store | 7 (7.2%) |  |
| Travel | 6 (6.2%) |  |
| Clear known contact | 67 (69.1%) |  |

More than half of participants (56%) worked in a high-risk occupational for COVID-19 infection. Of these, many patients were front-line health care workers including 1 physician (1.0%), 11 nurses (11.3%), 13 personal support workers (13.4%), 5 shelter workers (5.1%) and 6 who worked in cleaning or environmental services in a health care setting (6.2%).

### Feasibility and Health Care Utilization

Across the 97 patients, 415 visits took place with a family physician or nurse; 62% were booked as video visits and 38% were booked as phone visits. **Table 2** documents visit type and utilization of the program. The median time from viral swab positive test to first COVIDcare@Home assessment was 3 days (IQR 2). The median virtual length of stay in the program was 8 days (IQR 5), with an average of 4.4 visits (SD 2.5) per patient.

**Table 2:** COVIDCare@Home Process Measures.

|  | Value | Interquartile range or Standard deviation |
| --- | --- | --- |
| **Total number of visits** | 415 visits |  |
| Family Physician Staff/Resident | 251 visits (60.4%) |  |
| Registered Nurse | 164 visits (39.5%) |  |
| **Number of visits per patient (Average and SD)** | 4.4 visits | 2.5 (SD) |
| 1-2 visits | 24 patients (24.7%) |  |
| 3-5 visits | 51 patients (52.6%) |  |
| 6-8 visits | 13 patients (13.4%) |  |
| 9-13 visits | 9 patients (9.3%) |  |
| **\*Time from swab results to first visit (Median & IQR)** | 3 days | 2 (IQR) |
| **Virtual Length of stay in program in days (Median & IQR)** | 8 days | 5 (IQR) |

\*n=94, 3 patients were not swabbed.

Of the 97 patients, 5 (5.1%) required escalation with an in-person visit to the AACU or ED. 16% of patients required consultation with a social worker. **Table 3** outlines the health care utilization within the program.

**Table 3:** General program utilization and delivery.

|  |  |
| --- | --- |
|  | Number(%) of patients |
| **Sent a(n):** |  |
| Oximeter | 24 (24.5%) |
| Thermometer (n = X eligible) | 5 |
| **Health service utilization:** |  |
| **Virtual General Internal Medicine** (reasons below) | 4 (4.1%) |
| Worsening symptoms | 2 (2.1%) |
| Co-morbidity management | 1 (1.0%) |
| Rule out other disease | 1 (1.0%) |
| **Social Worker** (reasons below) | 16 (16.5%) |
| Find PCP | 9 (9.3%) |
| Financial or food insecurity | 6 (6.2%) |
| Mental health | 4 (4.1%) |
| **Pharmacy** | 6 (6.2%) |
| **Acute Ambulatory Care Unit** | 1 (1.0%) |
| **Emergency Department** | 4 (4.2%) |
| Self-Referred | 2 (2.1%) |
| Referred through program | 2 (2.1%) |
| **Hospitalization** | 0 (0%) |

## Interpretation

Analysis of the first 97 patients in the COVIDCare@Home program demonstrates that a team-based, family medicine-led remote monitoring program is a feasible and safe option to manage COVID-19 patients in the community. The median virtual length of stay of 8 days in the program and an average of 4.4 visits per patient suggest strong patient retention over the typical time course of COVID-19. Preliminary analysis of health services utilization shows limited use of acute care services, including no hospitalizations. In comparison, as of July 2, 2020, 15% of patients with COVID-19 across Canada were hospitalized with 20% being 40-50 years old (1). These results suggest that the COVIDCare@Home model may help limit the burden of COVID-19 on acute care settings and improve the system level response to the pandemic.

Six similar remote monitoring programs have been described in the literature, including three from the United States, and one each from Australia, China, and Canada (5,6,13,14,16,17). In terms of program design, our model was intermediate in its intensity. On the low intensity end, a model from Minneapolis used existing processes for post-surgical remote monitoring that were easy to scale: scrolling newsfeed with reminders, daily symptom questionnaires, an option to send questions, and a dashboard to monitored over 1300 patients (14). On the high end, the Australian model had one nurse per 25 patients per shift, and checked monitoring data three times a day, with 2 phone or video check-ins, and everyone received a pulse oximeter (16). For programs with similar patient populations to COVIDCare@Home, the ED and hospital admission rates were all relatively low (1-12%), however direct comparison is challenging due to the different data reporting approaches.

Our model was family medicine-led, while other programs were led by specialists or a mix of clinicians from different disciplines (5,6,13,17). Initial results suggest this approach is well-suited to managing the wide range of medical symptoms and comorbidities prevalent in COVID-19 patients. Additional support by a specialist was required in a few cases and often could be provided virtually. Of note, our model linked patients to their own PCP to maintain continuity of care, which was only mentioned by one other program (5). While 77% of patients reported having a PCP, many stayed in the program as it filled a gap in services during the pandemic. First, the COVIDCare@Home program could provide daily follow-up, which was impractical for many community primary care physicians. Second, having a dedicated interprofessional team with expertise in COVID-19 may have been reassuring to patients and their PCP. Future work should explore how to best support the PCP-patient relationship in remote monitoring programs.

Our team-based model and primary care expertise enabled us to support the mental health and social needs of patients. This was particularly relevant as 24.7% of our patients belonged to occupational groups (personal support workers, shelter workers and cleaners) who are more likely to contract COVID-19, and more likely to have social issues that increase the risk of poor health outcomes (18). For example, personal support workers are likely to be racialized, women, and have precarious employment arrangements (19). This aligns with several studies demonstrating that women, people of colour, and recent immigrants have higher rates of COVID-19 and worse outcomes (20–23). A quarter of our patients did not have a regular PCP, which may reflect barriers including language, lack of local physicians and difficulty navigating the health system (24,25). While digital health or virtual care can raise concerns about increasing health disparities among groups, the COVIDCare@Home model suggests that in certain cases, virtual care may improve access to those who are not well served by the healthcare system. In this case, patients were able to access social workers/mental health providers at no cost, sometimes for critical resources such as access to food. Other published remote monitoring program for COVID-19 did not include resources to support mental health or address the social determinants of health, though some planned to do so (5,16). Mental health support and addressing the social determinants of health is a key part of the care of people with COVID-19, to support them in maintaining quarantine and ensure better outcomes.

### Limitations

While this study adds important insights to the growing literature on remote monitoring for patients with COVID-19 and more generally, there are several limitations. We have examined the first 97 patients, who were enrolled in the first 5 weeks of the program, and longer-term data collection with higher numbers will certainly yield more lessons. Further, the sample population was biased by the local COVID-19 testing prioritization at the time (focusing on health care workers and those from high-risk congregate living situations) (1). Older patients who lived in long-term care settings were not included in the program, as there were other services available to support their unique health needs. Flexible use of phone or video generally enabled broad access by patients, but those without access to a phone could not participate in the program. The initial model was fairly resource intensive with 7-day a week coverage from all team members. In the future, we can expect that there will be fluctuations in need with periodic outbreaks, so the use of electronic surveys, remote-monitoring apps, automated dashboards and greater integration of the caregiver role may make it easier to rapidly adjust capacity. This study did not include a control group to directly measure efficacy, but a detailed programmatic evaluation is underway to quantify the impact of COVIDCare@Home.

## Conclusions

This program showed that a multidisciplinary, family medicine-led, remote monitoring program for COVID-19 is safe and feasible. The primary care model may be more adaptable to evolving patient and system needs, and easier to replicate in settings with limited access to specialty care. Given that certain populations are disproportionally impacted by COVID, remote monitoring programs should consider how to improve health equity through increased virtual support to address social determinants of health. Virtual care approaches like COVIDCare@Home that limit unnecessary hospitalizations may be essential as we head into a second wave.

## Data Availability

Portions of the data are available upon request to the corresponding author.

## Supplement 1

COVID-19 Clinical Assessment Tip Sheet

## Data Sharing Statement

Portions of the data are available upon request to the corresponding author.

## Acknowledgement

The authors would like to thank the full team of clinicians and partners who developed and continue to deliver the COVIDCare@Home program. Further thanks to the patients who participated in this program.

## Contributors Statement

PA led on all aspects of this program and manuscript. GM supported the conception of the study and supervised the overall design and activities of the manuscript. CL contributed to the evaluation of the program and the comparison to other literature. SC contributed to the evaluation of the program and supported the data analysis. NP, RH, RS, EG supported the conception and development of the program and development of the paper. SB, OB, and DM provide oversight to the program development and delivery and supported manuscript conceptualization and writing. All authors read, edited and approved the final manuscript prior to publication.

## References

1. Government of Canada. Epidemiological summary of COVID-19 cases in Canada: Hospitalizations, intensive care unit (ICU) admissions and deaths. 2020. Available from: https://health-infobase.canada.ca/covid-19/epidemiological-summary-covid-19-cases.html#a7. Accessed July 3, 2020.

2. Greenhalgh T, Koh GCH, Car J. Covid-19: A remote assessment in primary care.BMJ. 2020 Mar 25;368.

3. Berlin DA, Gulick RM, Martinez FJ. Severe Covid-19. N Engl J Med. 2020 May 15;

4. Gandhi RT, Lynch JB, del Rio C. Mild or Moderate Covid-19. N Engl J Med. 2020 Apr 24;

5. Medina M, Babiuch C, Card M, Gavrilescu R, Zafirau W, Boose E, et al. Home monitoring for COVID-19. Cleve Clin J Med. 2020 May 14;

6. Lam PW, Sehgal P, Andany N, Mubareka S, Simor AE, Ozaldin O, et al. A virtual care program for outpatients diagnosed with COVID-19: a feasibility study. CMAJ Open. 2020 Apr 23;8(2):E407–13.

7. Argenziano MG, Bruce SL, Slater CL, Tiao JR, Baldwin MR, Graham Barr R, et al. Characterization and clinical course of 1000 patients with coronavirus disease 2019 in New York: retrospective case series. BMJ. 2020;369(m1996).

8. Song JY, Yun JG, Noh JY, Cheong HJ, Kim WJ. Covid-19 in South Korea -Challenges of subclinical manifestations. N Engl J Med. 2020 May 7;382(19):1858–9.

9. Noh JY, Yoon JG, Seong H, Choi WS, Sohn JW, Cheong HJ, et al. Asymptomatic infection and atypical manifestations of COVID-19: Comparison of viral shedding duration. J Infect. 2020;

10. Saeedi N &, Naderpour Z, Saeedi M. Primer on COVID-19 for Clinicians: Clinical Manifestation and Natural Course. Adv J Emerg Med. 2020 May 4;4(2s):62.

11. Majeed A, Maile EJ, Bindman AB. The primary care response to COVID-19 in England’s National Health Service. J R Soc Med. 2020 Jun 1;113(6):208–10.

12. Williams S, Tsiligianni I. COVID-19 poses novel challenges for global primary care. npj Prim Care Respir Med. 2020 Dec 1;30(1):1–3.

13. Kricke G, Roemer PE, Barnard C, Devin J, Henschen BL, Bierman JA, et al. Rapid Implementation of an Outpatient Covid-19 Monitoring Program. New Engl J Med Catal. 2020;

14. Annis T, Pleasants S, HultmanGretchen Lindemann E, Thompson JA, Billecke S, et al. Rapid Implementation of a COVID-19 Remote Patient Monitoring Program. J Am Med Inf Assoc. 2020.

15. Centers for Disease Control and Prevention. Interim Clinical Guidance for Management of Patients with Confirmed Coronavirus Disease (COVID-19). 2020.

16. Hutchings O, Dearing C, Jagers D, Shaw M, Raffan F, Jones A, et al. Virtual health care for community management of patients with COVID-19. Pre-Print. 2020 May 13;

17. Xu H, Huang S, Qiu C, Liu S, Deng J, Jiao B, et al. Monitoring and Management of Home-Quarantined Patients With COVID-19 Using a WeChat-Based Telemedicine System: Retrospective Cohort Study. J Med Internet Res. 2020 Jul 2;22(7):e19514.

18. Toronto Public Health. COVID-19 and the Social Determinants of Health: What do we know? Toronto; 2020.

19. Van Houtven CH, DePasquale N, Coe NB. Essential Long-Term Care Workers Commonly Hold Second Jobs and Double-or Triple-Duty Caregiving Roles. J Am Geriatr Soc. 2020 May 14;jgs.16509.

20. Laurencin CT, McClinton A. The COVID-19 Pandemic: a Call to Action to Identify and Address Racial and Ethnic Disparities. J Racial Ethn Heal Disparities. 2020 Jun 1;7(3):398–402.

21. Kantamneni N. The impact of the COVID-19 pandemic on marginalized populations in the United States: A research agenda. Vol. 119, Journal of Vocational Behavior. Academic Press Inc., 2020.

22. Centers for Disease Control and Prevention. COVID-19 in Racial and Ethnic Minority Groups. 2020.

23. Garg S, Kim L, Whitaker M, O’Halloran A, Cummings C, Holstein R, et al. Review of “Hospitalization rates and characteristics of patients hospitalized with laboratory-confirmed coronavirus disease 2019 — COVID-NET, 14 states, March 1–30, 2020. 2020.

24. Glazier R, Gozdyra P, Kim M, Bai L, Kopp A, Schultz S, et al. Geographic Variation in Primary Care Need, Service Use and Providers in Ontario, 2015/16. Ontario; 2018.

25. Ahmed S, Shommu NS, Rumana N, Barron GRS, Wicklum S, Turin TC. Barriers to Access of Primary Healthcare by Immigrant Populations in Canada: A Literature Review. J Immigr Minor Heal. 2016 Dec 1;18(6):1522–40.

